## Supplementary Table 1 and Supplementary Figures 1-5 for "Distorted TCR repertoires define multisystem inflammatory syndrome in children"

**Supplementary Table 1: TCR repertoire metrics of samples**

| Sample | Patient | Days post COVID19 diagnosis | Sample Type | Clones | Reads | Shannon Evenness |
|---|---|---|---|---|---|---|
| 10 | 10 | 8 | severe COVID-19 | 420 | 44275 | 0.93 |
| 11 | 11 | 14 | severe COVID-19 | 1147 | 35585 | 0.94 |
| 12_1 | 12 | 30 | mild COVID-19 | 157 | 25413 | 0.93 |
| 12_2 | 12 | 80 | mild COVID-19 | 55 | 41886 | 0.82 |
| 13_1 | 13 | 7 | mild COVID-19 | 86 | 15440 | 0.9 |
| 13_2 | 13 | 29 | mild COVID-19 | 156 | 17239 | 0.92 |
| 13_3 | 13 | 46 | mild COVID-19 | 101 | 14250 | 0.86 |
| 13_4 | 13 | 56 | mild COVID-19 | 535 | 85494 | 0.9 |
| 14 | 14 | 2 | mild COVID-19 | 497 | 27125 | 0.93 |
| 15 | 15 | 12 | severe COVID-19 | 503 | 39541 | 0.94 |
| 16 | 16 | 8 | severe COVID-19 | 472 | 30621 | 0.94 |
| 18_1 | 18 | 7 | mild COVID-19 | 1837 | 41922 | 0.94 |
| 18_2 | 18 | 8 | mild COVID-19 | 1585 | 58978 | 0.93 |

| | | | | | | |
|---|---|---|---|---|---|---|
| 18_3 | 18 | 9 | mild COVID-19 | 1876 | 29961 | 0.94 |
| 19 | 19 | 17 | severe COVID-19 | 2135 | 113638 | 0.95 |
| 2 | 2 | 0 | mild COVID-19 | 897 | 61748 | 0.91 |
| 20_1 | 20 | 13 | severe COVID-19 | 5124 | 69895 | 0.95 |
| 20_2 | 20 | 17 | severe COVID-19 | 1085 | 157191 | 0.9 |
| 21 | 21 | 9 | mild COVID-19 | 2645 | 90117 | 0.94 |
| 22_1 | 22 | 66 | mild COVID-19 | 1387 | 86362 | 0.92 |
| 22_2 | 22 | 77 | mild COVID-19 | 3161 | 123745 | 0.92 |
| 3 | 3 | 13 | severe COVID-19 | 706 | 54406 | 0.93 |
| 4 | 4 | 5 | severe COVID-19 | 1326 | 73412 | 0.95 |
| 5 | 5 | 17 | severe COVID-19 | 660 | 50959 | 0.92 |
| 6 | 6 | 9 | severe COVID-19 | 1354 | 78079 | 0.95 |
| 7 | 7 | 7 | severe COVID-19 | 1095 | 64906 | 0.95 |
| 8 | 8 | 5 | severe COVID-19 | 361 | 19634 | 0.94 |
| 9_1 | 9 | 24 | mild COVID-19 | 244 | 25617 | 0.93 |

| | | | | | | |
|---|---|---|---|---|---|---|
| 9_2 | 9 | 52 | mild COVID-19 | 904 | 66203 | 0.92 |
| 9_3 | 9 | 69 | mild COVID-19 | 825 | 149883 | 0.92 |
| 9_4 | 9 | 79 | mild COVID-19 | 1066 | 81956 | 0.92 |
| H10 | H10 | NA | pre-COVID-19 | 6921 | 142983 | 0.94 |
| H12 | H12 | NA | pre-COVID-19 | 9529 | 117661 | 0.94 |
| H15 | H15 | NA | pre-COVID-19 | 5801 | 103000 | 0.93 |
| H17 | H17 | NA | pre-COVID-19 | 3885 | 74079 | 0.93 |
| H18 | H18 | NA | pre-COVID-19 | 7134 | 92026 | 0.93 |
| H30 | H30 | NA | pre-COVID-19 | 4214 | 65193 | 0.94 |
| H31 | H31 | NA | pre-COVID-19 | 806 | 109028 | 0.93 |

**Supplementary figure legends**

**Supplementary Figure 1**: a) The fraction of TRBV11-2 chains in patient repertoires as a function of the patients' age with Spearman's Rank Correlation coefficient (ρ) and P value. b) The fraction of TRBV11-2 chains in patient repertoires as a function of the number of days after COVID-19 diagnosis with Spearman's Rank Correlation coefficient (ρ) and P value. c) The fraction of TRBV11-2 chains in the repertoires of antibody negative and positive patients. Significance determined by unpaired Wilcoxon test between each paediatric group, with adjustment for multiple comparisons using Benjamini-Hochberg correction, indicated by: * p<0.05, ** p<0.01, and *** p<0.001. Lack of notation for specified comparisons indicates no statistical significance.

**Supplementary Figure 2**: a) The number of clones/million reads in a repertoire as a function of the patients' age with Spearman's Rank Correlation coefficient (ρ) and P value.

b) The number of clones/million reads in a repertoire as a function of the number of days after COVID-19 diagnosis with Spearman's Rank Correlation coefficient (ρ) and P value.

c) The number of clones/million reads in the repertoires of patients with or without co-morbidities. Significance determined by unpaired Wilcoxon test between each paediatric group, with adjustment for multiple comparisons using Benjamini-Hochberg correction, indicated by: * p<0.05, ** p<0.01, and *** p<0.001. Lack of notation for specified comparisons indicates no statistical significance.

**Supplementary Figure 3:** a) The fraction of class I MIRA clones in the repertoires of patient cohorts. Significance determined by unpaired Wilcoxon test between each paediatric group, with adjustment for multiple comparisons using Benjamini-Hochberg correction, indicated by: * p<0.05, ** p<0.01, and *** p<0.001. Lack of notation for specified comparisons indicates no statistical significance. b) The number of class I MIRA clones as the function of the total number of clones in the patient repertoires with Spearman's Rank Correlation coefficient (ρ) and P value. c) The fraction of class II MIRA clones in the repertoires of patient cohorts. Significance determined by unpaired Wilcoxon test between each paediatric group, with adjustment for multiple comparisons using Benjamini-Hochberg correction, indicated by: * p<0.05, ** p<0.01, and *** p<0.001. Lack of notation for specified comparisons indicates no statistical significance. d) The number of class II MIRA clones as the function of the total number of clones in the patient repertoires with Spearman's Rank Correlation coefficient (ρ) and P value.

**Supplementary Figure 4**: Distance to class I MIRA distribution of clones in the repertoires of children with mild or severe COVID-19. Distance to class I MIRA distributions were fitted with Gaussian probability density functions and the difference between patient cohorts was assessed by comparing a model fitting all symptom groups together to a model taking into account the differences between symptom groups. The difference between the two models was determined with ANOVA analysis, with all p-values corrected for multiple hypothesis testing using Benjamini-Hochberg adjustment. Solid curves show the probability distribution functions determined by fitting the patient cohorts separately, shaded areas show 75% confidence intervals. P-values denote the statistical assessment of the models fitting all patient cohorts together versus fitting each patient cohort separately. See the distance to class I MIRA distribution of TRBV11-2 clones in Figure 3b.

**Supplementary Figure 5**: Distance to class II MIRA distribution of clones in the repertoires of children with mild or severe COVID-19. Distance to class II MIRA distributions were fitted with Gaussian probability density functions and the difference between patient cohorts was assessed by comparing a model fitting all symptom groups together to a model taking into account the differences between symptom groups. The difference between the two models was determined with ANOVA analysis, with all p-values corrected for multiple hypothesis testing using Benjamini-Hochberg adjustment. Solid curves show the probability distribution functions determined by fitting the patient cohorts separately, shaded areas show 75% confidence intervals. P-values denote the statistical assessment of the models fitting all patient cohorts together versus fitting each patient cohort separately. See the distance to class II MIRA distribution of TRBV11-2 clones in Figure 3c.

#### Supplementary Figure 1

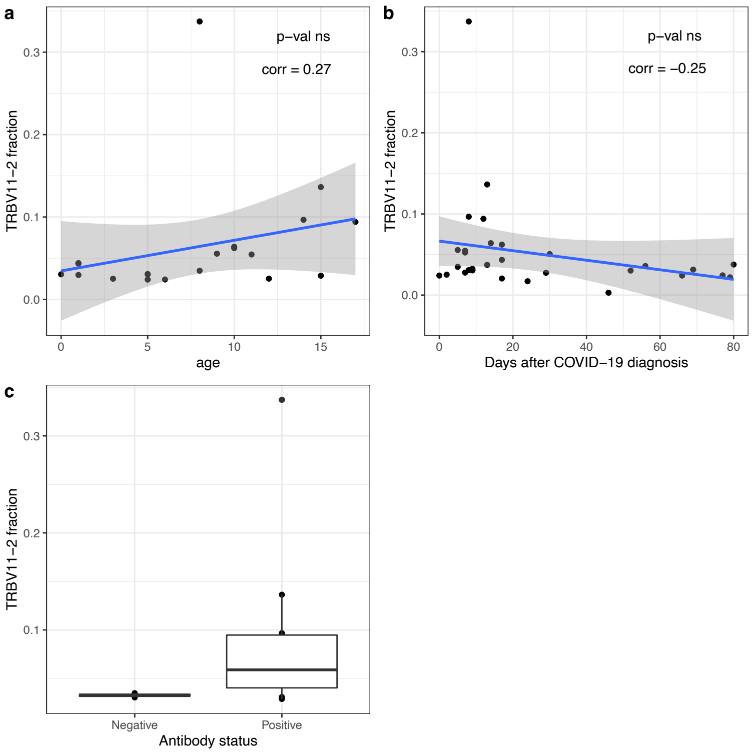

#### Supplementary Figure 2

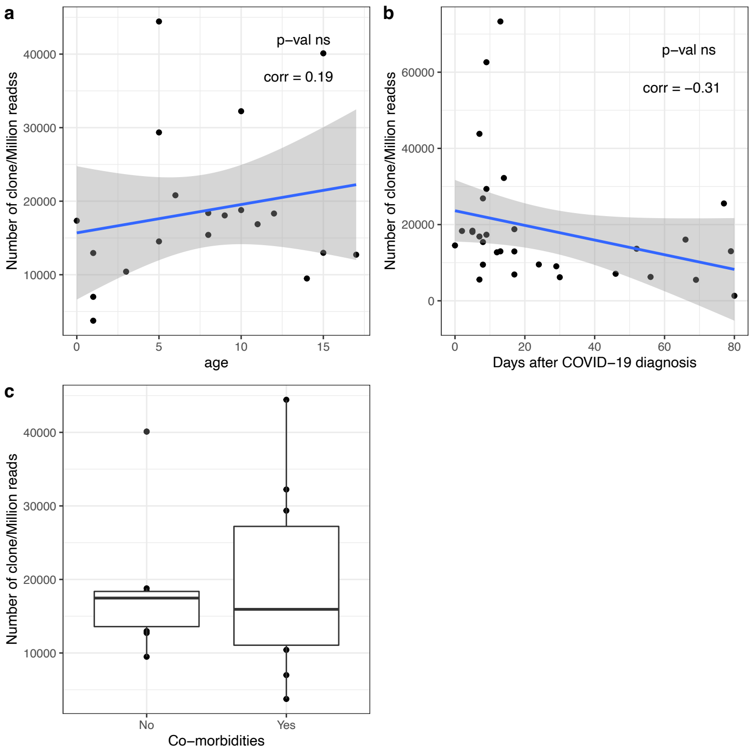

### Supplementary Figure 3

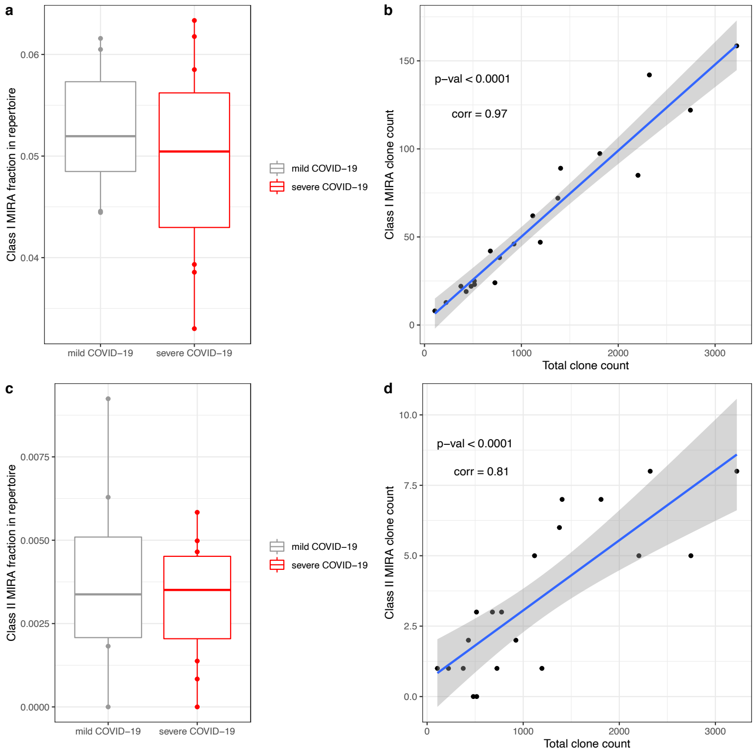

### Supplementary Figure 4

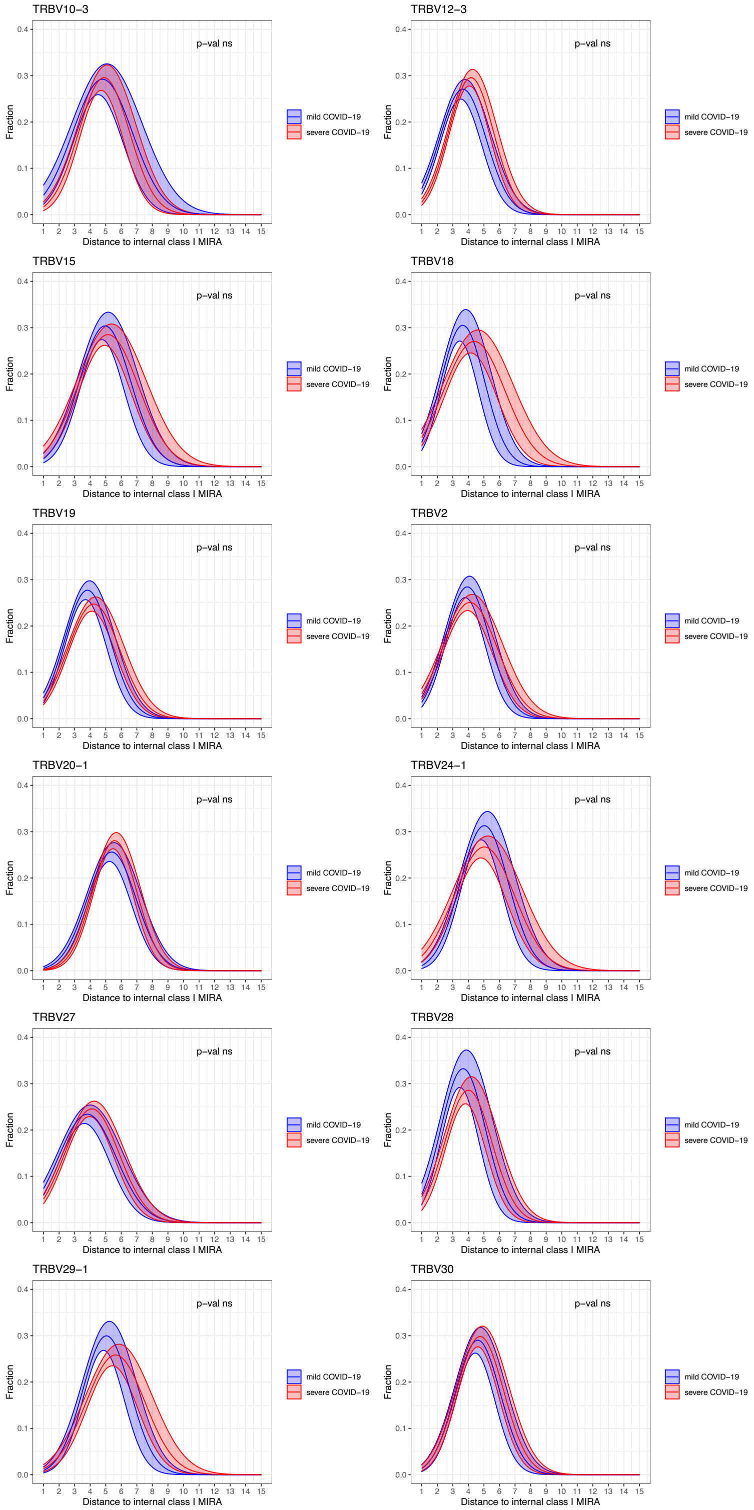

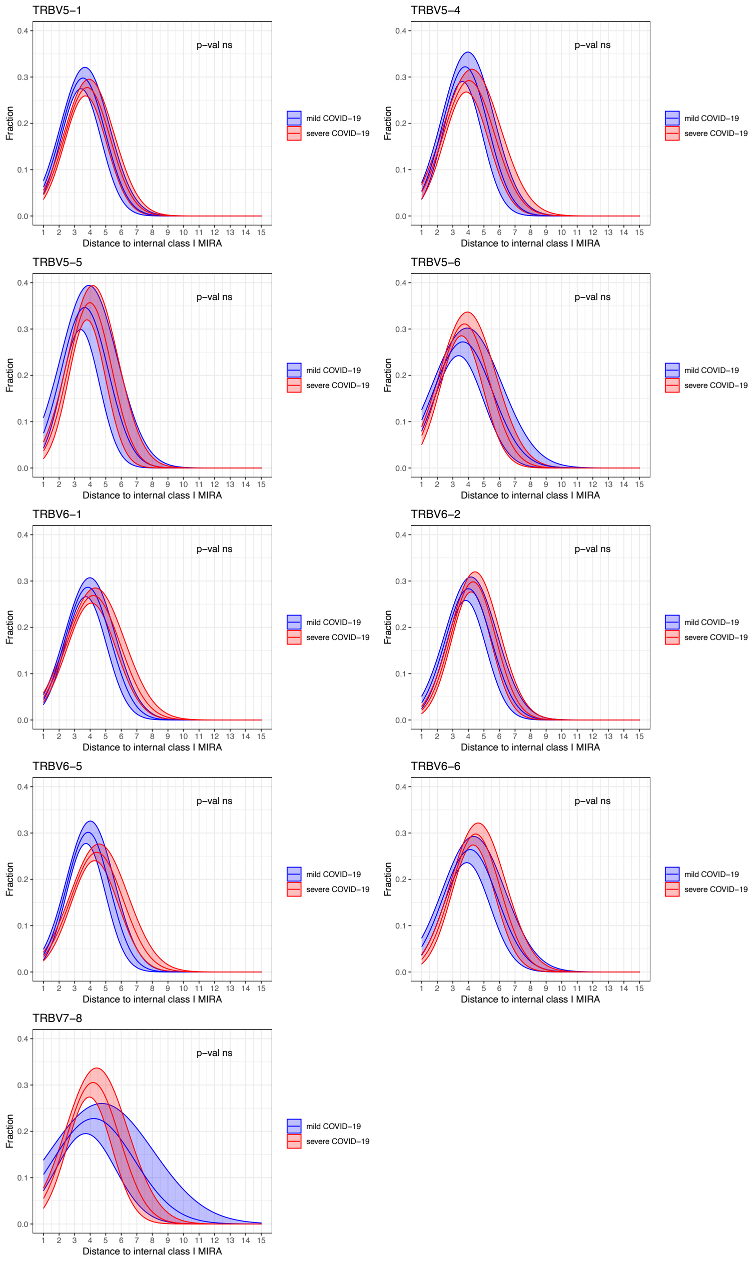

TRBV5−1
p−val ns
Fraction
0.0
0.1
0.2
0.3
0.4
1 2 3 4 5 6 7 8 9 10 11 12 13 14 15
Distance to internal class I MIRA
mild COVID−19
severe COVID−19
TRBV5−4
p−val ns
Fraction
Distance to internal class I MIRA
mild COVID−19
severe COVID−19
TRBV5−5
p−val ns
Fraction
Distance to internal class I MIRA
mild COVID−19
severe COVID−19
TRBV5−6
p−val ns
Fraction
Distance to internal class I MIRA
mild COVID−19
severe COVID−19
TRBV6−1
p−val ns
Fraction
Distance to internal class I MIRA
mild COVID−19
severe COVID−19
TRBV6−2
p−val ns
Fraction
Distance to internal class I MIRA
mild COVID−19
severe COVID−19
TRBV6−5
p−val ns
Fraction
Distance to internal class I MIRA
mild COVID−19
severe COVID−19
TRBV6−6
p−val ns
Fraction
Distance to internal class I MIRA
mild COVID−19
severe COVID−19
TRBV7−8
p−val ns
Fraction
Distance to internal class I MIRA
mild COVID−19
severe COVID−19

### Supplementary Figure 5

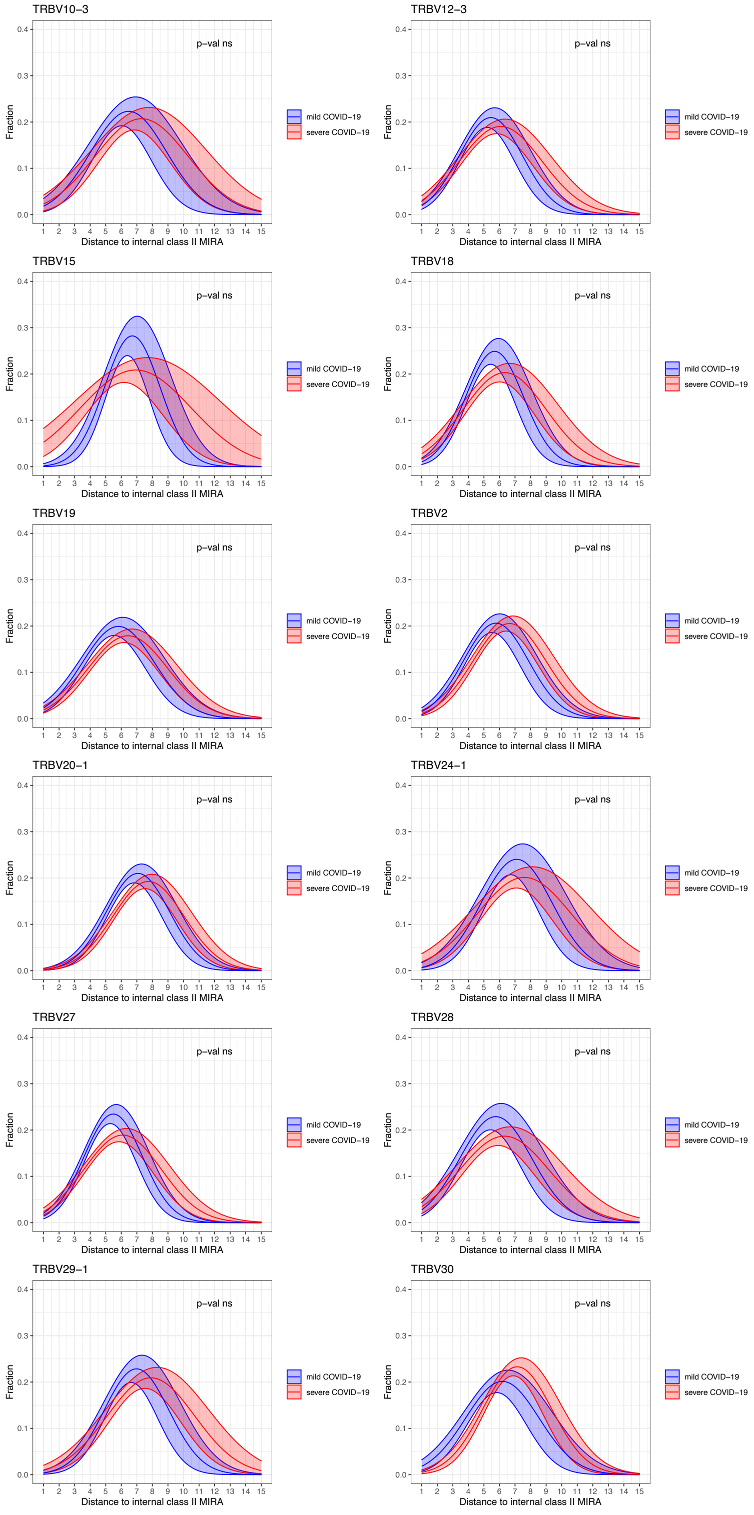

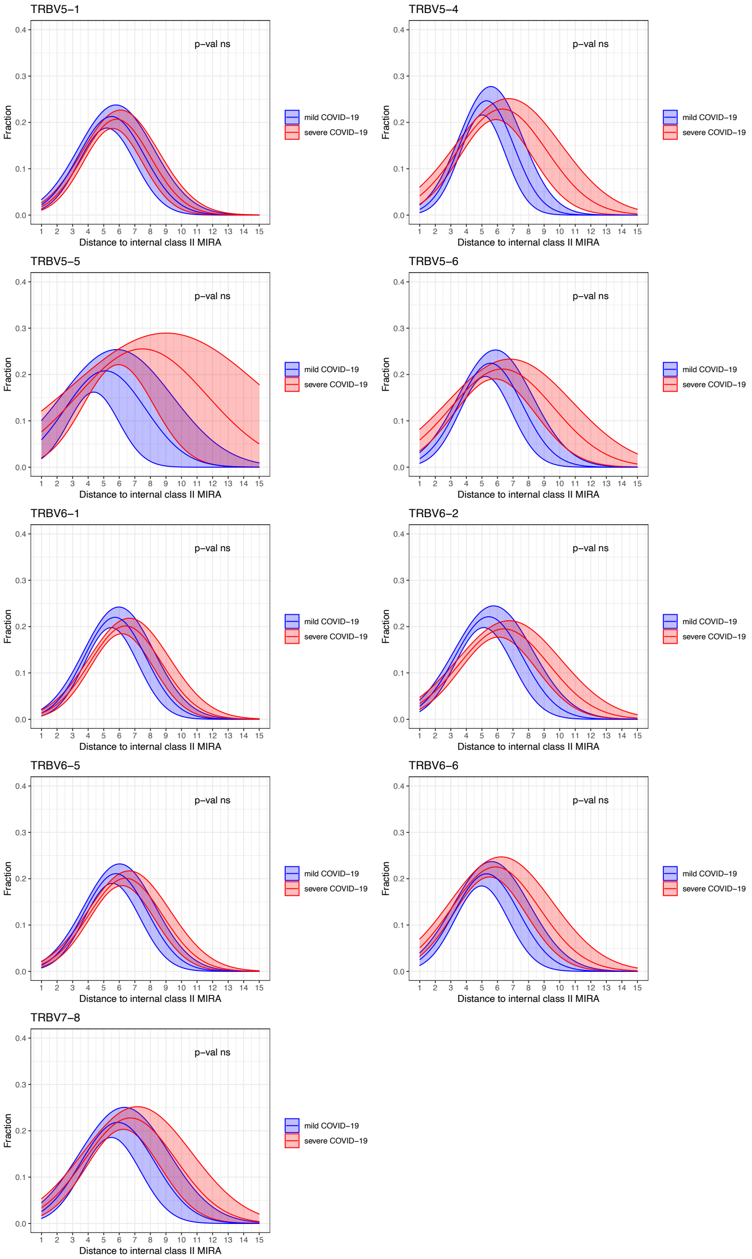

TRBV5−1
TRBV5−4
TRBV5−5
TRBV5−6
TRBV6−1
TRBV6−2
TRBV6−5
TRBV6−6
TRBV7−8
p−val ns
Fraction
Distance to internal class II MIRA
mild COVID−19
severe COVID−19
